# Updated SARS-CoV-2 Single Nucleotide Variants and Mortality Association

**DOI:** 10.1101/2021.01.29.21250757

**Authors:** Shuyi Fang, Sheng Liu, Jikui Shen, Alex Z Lu, Yucheng Zhang, Kailing Li, Juli Liu, Lei Yang, Chang-Deng Hu, Jun Wan

## Abstract

Since its outbreak in December 2019, COVID-19 has caused 100,5844,555 cases and 2,167,313 deaths as of Jan 27, 2021. Comparing our previous study of SARS-CoV-2 single nucleotide variants (SNVs) before June 2020, we found out that the SNV clustering had changed considerably since June 2020. Apart from that the group SNVs represented by two non-synonymous mutations A23403G (S: D614G) and C14408T (ORF1ab: P4715L) became dominant and carried by over 95% genomes, a few emerging groups of SNVs were recognized with sharply increased monthly occurrence ratios up to 70% in November 2020. Further investigation revealed that several SNVs were strongly associated with the mortality, but they presented distinct distribution in specific countries, e.g., Brazil, USA, Saudi Arabia, India, and Italy. SNVs including G25088T, T25A, G29861T and G29864A were adopted in a regularized logistic regression model to predict the mortality status in Brazil with the AUC of 0.84. Protein structure analysis showed that the emerging subgroups of non-synonymous SNVs and those mortality-related ones in Brazil were located on protein surface area. The clashes in protein structure introduced by these mutations might in turn affect virus pathogenesis through conformation changes, leading to the difference in transmission and virulence. Particularly, we found that SNVs tended to occur in intrinsic disordered regions (IDRs) of Spike (S) and ORF1ab, suggesting a critical role of SNVs in protein IDRs to determine protein folding and immune evasion.

## Introduction

Been transmitted over one year, COVID-19 has caused 100,584,555 cases and 2,167,313 deaths worldwide as of Jan 27, 2021 (1). The global mortality rate of 2.15% greatly exceeded the estimated seasonal flu death rate in the USA, less than 0.1% according to the 2018-29 data from the US Centers for Disease Control and Prevention (CDC). Since its outbreak, diverse viral genomic mutations of SARS-CoV-2 have been observed (2, 3), including insertions, deletions, and single nucleotide variants (SNVs) (4, 5), which led to viral protein structure changes that potentially affect the transmission and virulence of COVID-19. SNVs have been extensively detected by massive and daily updated whole-genome sequencing. SARS-CoV-2 SNVs presented clustering characteristics (4) in terms of concurrence (5-8) that is likely to be linked to the complex mechanism of epistatic gene interactions in COVID-19 viral evolution. Therefore, timely updates of SARS-CoV-2 mutations, especially critical SARS-CoV-2 mutations that are associated with COVID-19 mortality rate, become a necessary and important step in the fight against COVID-19. However, such investigations remain scarce due to limited clinical information along with whole-genome sequences (9-11).

Following our previous report (5), we analyzed a total of 146,045 SARS-CoV-2 high-quality complete genomes downloaded from GISAID after June 1, 2020 (12). The majority of SARS-CoV-2 genomes have evolved with a dominant SNV cluster represented by non-synonymous mutations A23403G (S: D614G) and C14408T (ORF1ab: P4715L), in addition to C241T at the upstream of ORF1ab and another synonymous mutation C3037T on ORF1ab. According to two-way clustering analysis on SNVs harbored by over 1% of SARS-CoV-2 genomes, additional SNVs were uncovered with increasing occurrence ratios over recent months. Even though two dominant amino acid (AA) changes, ORF1ab: 4715L and S: 614G variants, were reported to be strongly correlated with mortality (9), their death rates have been relatively stable. Or at least they were not strongly correlated with occurrence ratios of these two mutations which were carried by over 99% of patients now. Therefore, we did a systematic analysis on geographical distributions of SNVs and their corresponding mortality rates. By performing enrichment analysis on 6,845 SARS-CoV-2 genomes with clinical information, we identified multiple SNVs that were significantly related to COVID-19-associated death. In the meantime, we did protein structure analysis on non-synonymous SNVs showed clashes caused upon mutations, in turn, might contribute to viral transmission and mortality. Some SNVs were also found with tendency of occurrence in intrinsic disordered regions (IDRs) of S and ORF1ab. Our findings can help to better understand the pathogenesis of SARS-CoV-2 at the genetic level, possibly providing insights into therapeutic intervention and vaccine design in the future.

## Materials and Methods

### Data Collection

SARS-CoV-2 genome sequencing data and available clinical information were downloaded from GISAID from December 2019 to November 20^th^ 2020. 146,045 genomes with 1,748,802 SNVs along with 156,413 patient clinical information were collected for analysis. After filtering patients with the clinical status of “unknown” or “cryptic” for bias reduction, 6,845 genomes with clinical information were used for mortality analysis.

### Enrichment analysis

Candidate SNVs related to mortality were first estimated based on their occurrence rates in the death and non-death group. Enrichment analysis was conducted for each SNV. Fold Change (FC) of one SNV was calculated as the ratio of the SNV occurrence rate within the death group to that within the non-death group, whereas the statistical significance (*p*-value) was evaluated based on the hypergeometric distribution model (13).

### Regression Model

The elastic-net regularized logistic regression model (14) was employed to estimate the relationship between SNVs and mortality rate in individual countries. The elastic net method combines the penalties in Lasso (least absolute shrinkage and selection operator) and Ridge method, including the one with minimum mean cross-validated error as well as the other one resulting in the most regularized model. SNVs with |log_2_(FC)| > 1 and *p*-value < 0.05 based on the hypergeometric model were selected as features, while the clinical status (either death or non-death) of samples was set as the dependent variable for the regression model. To estimate the accuracy of the regression model, we took 5-fold cross-validation by using cv.glmnet function from the glmnet package in R (15) with setting the “family” option to “binomial”. The original samples were shuffled and randomly partitioned into 5 groups. We iteratively chose one group as the test data set for the model that was trained based on the rest 5 groups. The error of final model was generated based on the average of all 5 iterations. Considering the balance between accuracy and fewer number of SNVs, we finally chose the most regularized model given the value of λ that makes the cross-validated error within one standard error of the minimum (lambda.1se) (15).

### Protein Structure Analysis

We adopted PyMOL (16-18) to analyze and visualize protein structures for WT (Wuhan-Hu-1) and mutated proteins with identified non-synonymous SNVs. Mutagenesis tools in PyMOL were utilized to detect if a clash was generated upon the mutation. Properties of AAs were retrieved from the “Table of standard amino acid abbreviations and properties” on Wikipedia. The solved structures of S, nsp12, and nsp7 were downloaded from Protein Data Bank (PDB) (19): 6vyb for S using electron microscopy (20), 6m71 for nsp7, nsp12 using electron microscopy (21). Structures of other proteins/regions, e.g., incomplete regions of S, ORF3a, ORF10, and N, were predicted by the C-I-Tasser model (22-24): QHD43416 for S, QHD43417 for ORF3a, QHI42199 for ORF10, and QHD43423 for N.

## Results

### Emerging SNV clusters with temporal occurrence patterns

Previously, we discovered four major SNV groups according to the analysis of SNVs in genomes collected before June 2020 (5). Here we analyzed 81,042 more SARS-CoV-2 complete genomes from the GISAID database following the exclusion of low-coverage genomes. Same as our previous analysis (5), Wuhan-Hu-1 (NCBI Reference Sequence: NC_045512.2) was used as the reference genome to keep consistency. Approximately 5% of SARS-CoV-2 genomes were completely same with Wuhan-Hu1. In the remaining 95% genomes, a total of 20,477 nucleotide sites were identified to carry SNVs in at least one genome. The 52 SNVs occurring in greater than 3% of genomes were used for two-way clustering in the study.

The clustering pattern of SNVs after June 2020 (Figure 1A) were distinct from the one before June 2020 (5). Before June 2020, four major basically independent SNV groups were linked to the majority of SARS-CoV-2 genomes. It is not surprised to see that group A of SNVs has prevailed since June 2020, represented by A23403G (S: D614G) and C14408T (OFR1ab: P4715L) (5). This SNV group occupied over 99% of genomes that harbored at least one of 52 SNVs, confirming continuity of variant D614G on S protein (25-28). The results also indicated that SARS-CoV-2 evolution has reached a level by almost fully replacing these 4 nucleotides on the original strain, Wuhan-Hu-1. Among the 266 genomes without SNV group A, 246 carried one of the signature SNVs in group C, G11083T (ORF1ab: L3606F) (5). This suggested that ORF1ab: L3606F was mutually exclusive with S: D614G in this population with 66.3% genomes from Singapore. However, the other 95% (n = 4,211) of genomes with G11083T were dominated by instead of independent of group A of SNVs. Another representative SNV (G26144T) previously reported in group C (5) almost disappeared, being detected in only 53 genomes after June 2020. Meanwhile, Group B (T28144C, n = 199) and Group D (G1440A/G2891A, n = 7) (5) nearly diminished in newly collected SARS-CoV-2 genomes. Interestingly, most SNVs in Figure 1A were first identified in SARS-CoV-2 genomes collected before June 2020 (black boxes in Figure 1B), but their occurrence ratios gradually increased to higher levels (blue-filled boxes in Figure 1B) after several months. This might suggest potential incubation periods of SNVs during SARS-CoV-2 evolution.

**Figure 1.**
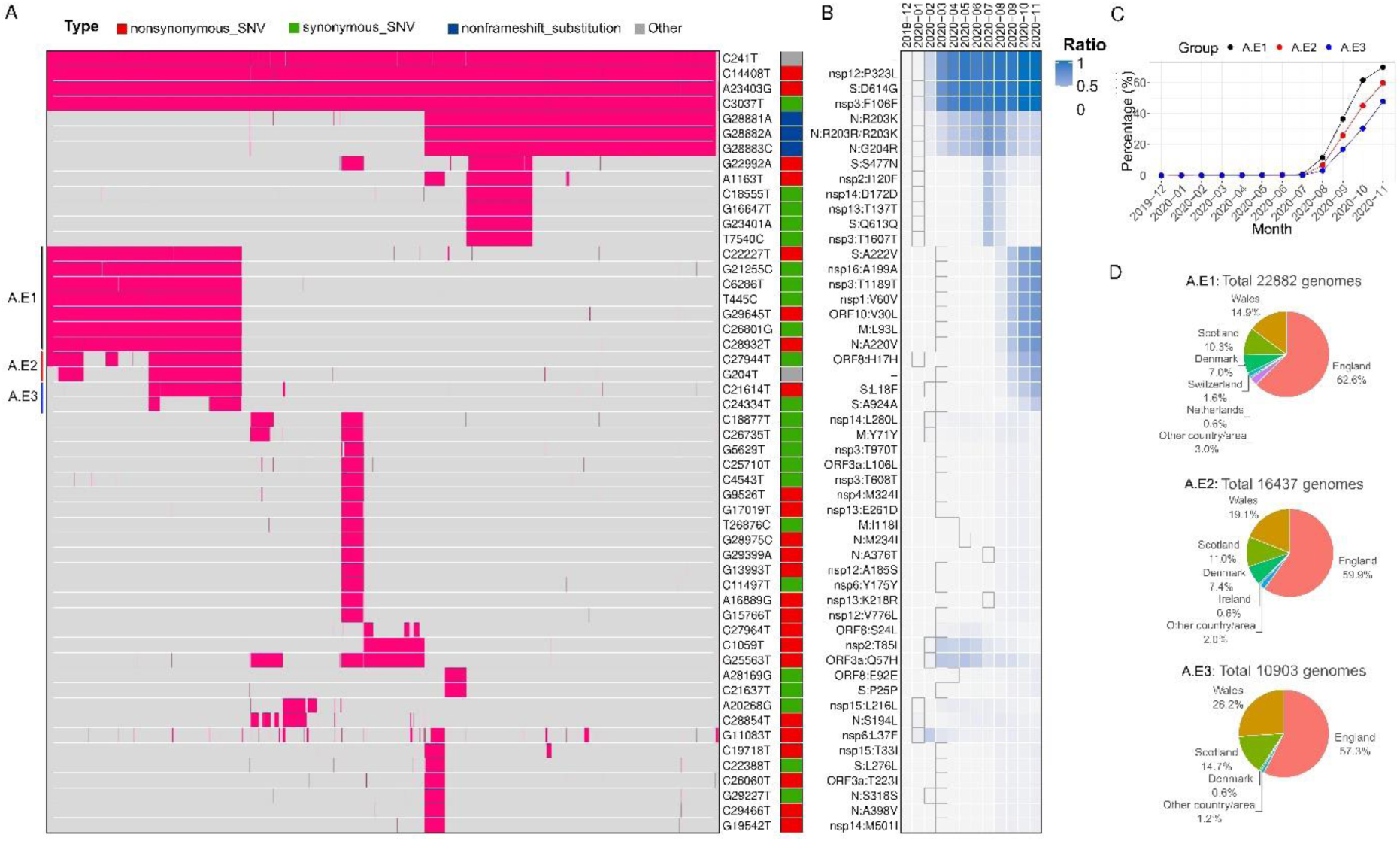
SNVs identified in more than 3% of SARS-CoV-2 genomes after June 1, 2020. (A) Two-way clustering of 52 high frequent SNVs with possible annotated amino acid (AA) changes in 76,926 genomes worldwide. (B) Monthly occurrence ratios of corresponding SNVs. (C) Temporal patterns of the emerging groups A.E1, A.E2, and A.E3. (D) Geographical distributions of emerging SNVs in groups A.E1-3, respectively.

A number of additional SNVs had become dominant most recently (Figures 1B and 1C). For example, group A.E1 included four other synonymous mutations (G21255C, T445C, C6286T, and C26801G), and three non-synonymous mutations, C22227T (S: A222V), G29645T (ORF10:V30L), C28932T (N: A220V), on proteins S, N, and ORF10, respectively. All of them were first identified in March 2020 in several different countries, e.g., Spain, England, Australia, and USA. This SNV group had gradually became dominant since the summer of 2020 (Figure 1C). Their occurrence ratios were significantly elevated from 10% in August to 70% in November. Almost 88% of 22,882 genomes with this group of SNVs (A.E1) were detected in England, Scotland, and Wales. The remaining 12% were identified in Denmark (7%), Switzerland (1.6%) and other counties (Figure 1D).

In group A.E2, synonymous SNV C27944T (ORF8:H17H) was first found in China in January 2020 and the other one G204T located at upstream of ORF1ab) was identified in USA in March 2020. Although C27944T on ORF8 is a synonymous mutation, it falls in a stem-loop structure of ORF8 which may influence ORF8 translation (29, 30). The A.E2 group had a very similar temporal pattern to that of A.E1, presenting high recent concurrence ratios (Figures 1C and 1D).

Two more SNVs on S, C21614T and C24334T, were identified in group A.E3. C21614T caused AA change on S: L18F, which was found lurking in human since February 2020 in England. The origin of C24334T can be traced back to Japan in March 2020. Similar to A.E1 and A.E2 groups, the occurrence ratio of A.E3 rose quickly since August 2020 (Figure 1C). They were carried by about 48% of new SARS-CoV-2 genomes collected in November. Although over 98% of these SNVs were found in England, we also observed 0.6% in Denmark.

### Mortality-associated SNVs

In order to investigate the association between SARS-CoV-2 SNVs and the mortality rate, we analyzed a total of 6,845 genomes with clinical information downloaded from GISAID. Among them, 665 samples (9.7%) were defined as death group with keywords “death”/”deceased”/”Hospitalized, deceased” in the clinical information, while the non-death group contains the genomes with all the other patient status. The monthly mortality rate kept an increasing tendency from December 2019 to April 2020 with a peak death ratio of 18.5% and mainly decreased in the following months (Figure 2A). The mortality rates seemed independent of the total number of collected patient samples.

**Figure 2.**
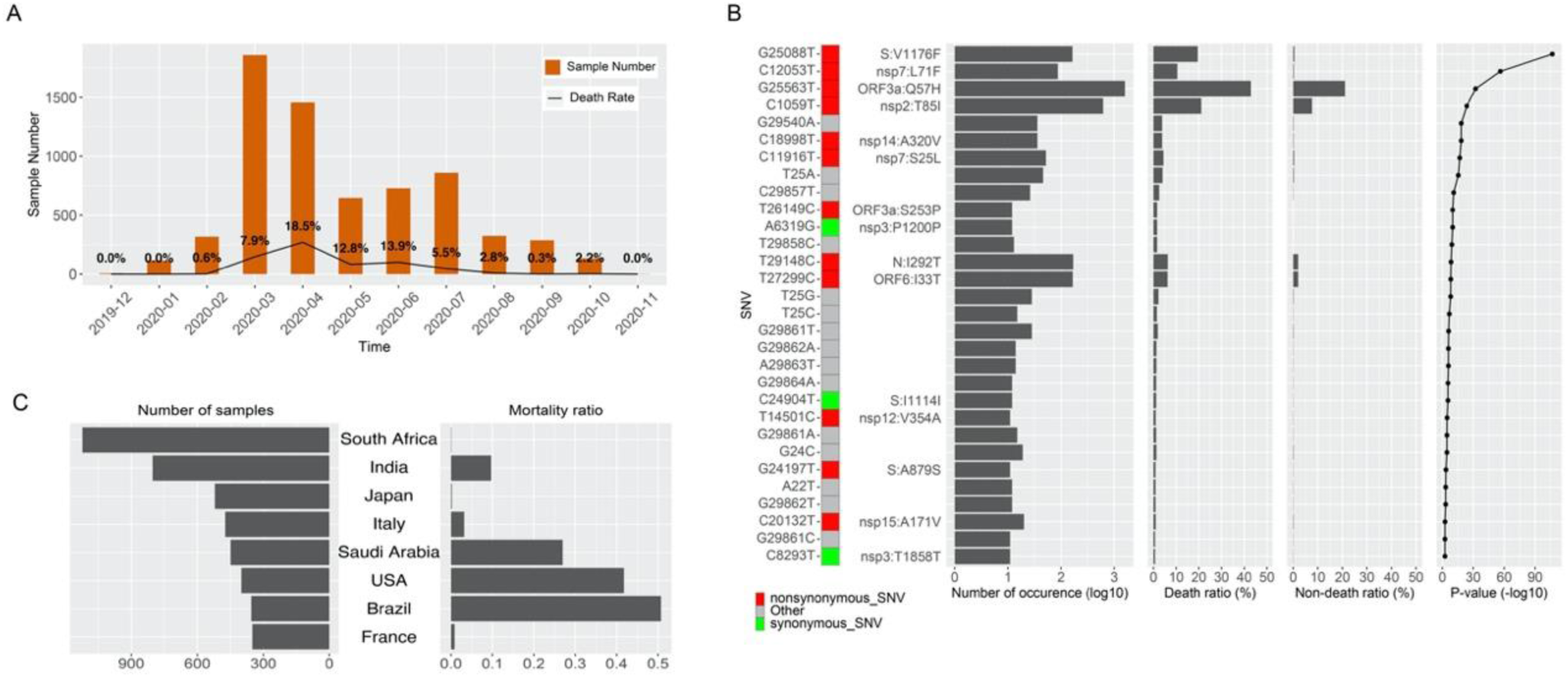
Morality related SNVs. (A) Numbers and ratios of SARS-CoV-2 genomes of the death group for each month. (B) 30 SNVs significantly over-represented in the death group with corresponding total numbers of occurrences, ratios of death and non-death groups, and enrichment *p*-value. (C) Number of samples and mortality rates for countries with at least 300 genomes collected.

By comparing SNV occurrence frequencies between the death and non-death groups, we identified 30 mortality related SNVs (Figure 2B). These SNVs were identified from at least 10 dead patients and significantly (*p* < 0.01) over-represented (FC > 2) in the death group (Figure 2B). However, strong sampling biases were observed in terms of death ratios of collected samples in individual countries (Figure 2C). For example, two of the top 3 countries that had the highest numbers of patients with clinical information came with very low death ratios, resulting in a diluted pool if they were included in the analysis along with all other countries. Hence, we decided to conduct similar enrichment analysis for 5 individual countries, e.g., Brazil, USA, Saudi Arabia, India, and Italy. These five countries had over 300 total patient samples for the death and non-death group. Using cutoffs of *p* < 0.01 and FC > 2, we found 12, 4, 5, 31, and 10 SNVs significantly over-represented in the death group in Brazil, USA, Saudi Arabia, India, and Italy separately (Figures 3A-3E).

**Figure 3.**
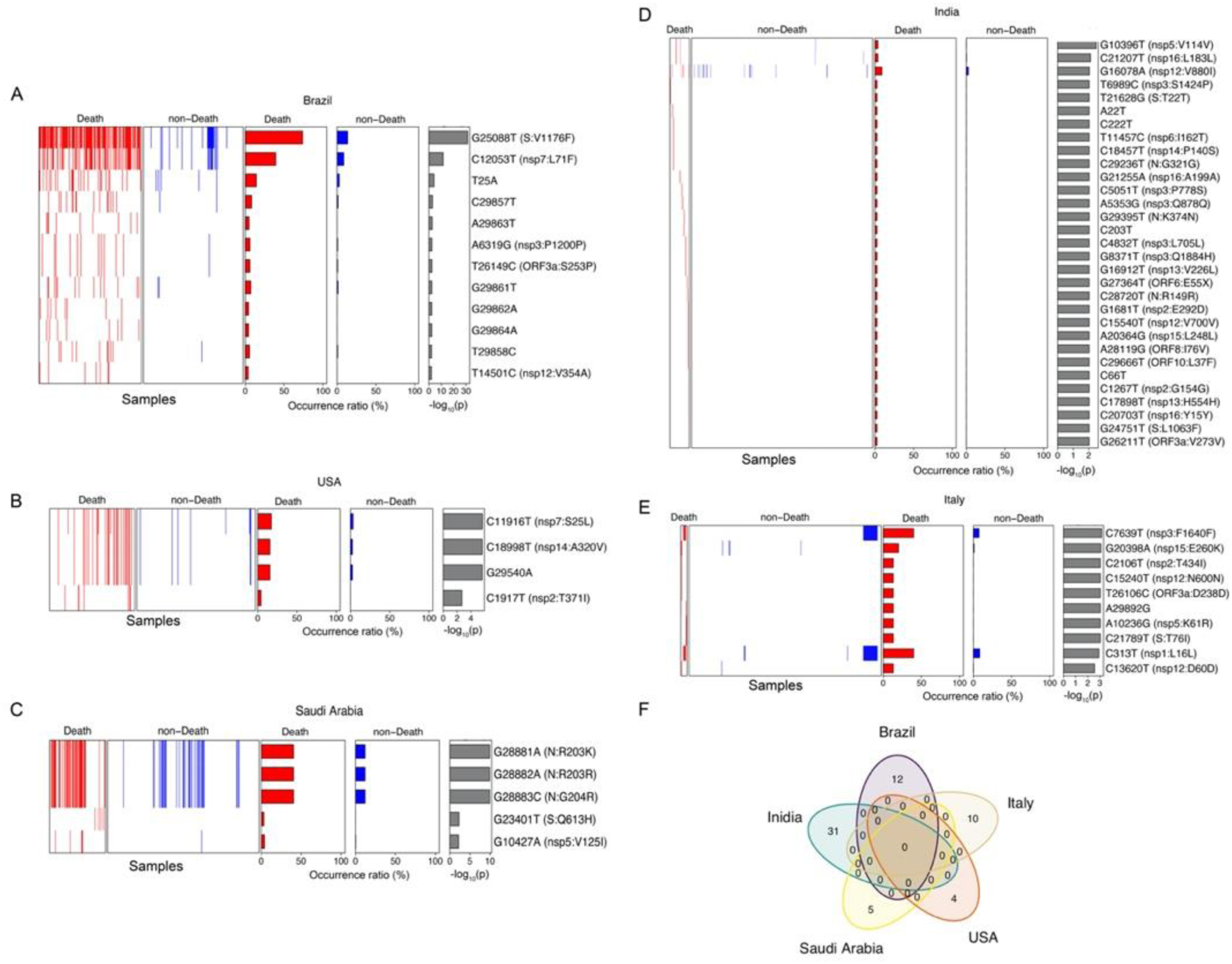
Mortality related SNVs identified in selected countries. (A) Brazil. (B) USA. (C) Saudi Arabia (D) India. (E) Italy. The red and blue colors represent the death and non-death groups, respectively. (F) Venn diagram of mortality related SNVs in these countries.

In Brazil, the top three SNVs includes two non-synonymous mutations: G25088T (S: V1176F) and C12053T (nsp7: L71F), together with T25A at upstream of ORF1a and ORF1ab (Figure 3A). About 73.9% of death cases carried G25088T while only 13.7% of non-death cases harbored this mutation (odds ratio of 17.80) in Brazil. Similar to G25088T, C12053T and T25A showed notable enrichment in the death group with odds ratios of 6.95 and 5.74, respectively. In USA (Figure 3B), 4 SNVs were significantly enriched in the death group (Figure 3D): C1917T (nsp2: T371I), C11916T (nsp7: S25L), C18998T (nsp14: A320A), and G29540A between N and ORF10. Three of them were carried by over 15% of death cases but by less than 5% of non-death cases, with odds ratio of 5.91, 6.97 and 6.90 for C11916T, C18998T and G29540A, respectively. These SNVs had high concurrence ratios among them, indicating strong functional collective correlations. 5 SNVs were identified to be significantly enriched in the death group in Saudi Arabia (Figure 3C), including three mutations at consecutive SARS-CoV-2 genome locations (28881-28883) on protein N, G28881A (N: R203K), G28882A (N: R203R), and G28883C (N: G204R). These three SNVs basically simultaneously occurred, covering 40.5% of genomes in the death group and 11.9% of non-death cases with odds ratio of 5.04. India is the country that has the second-largest number of COVID-19 patient samples following Brazil in our study pool. However, the numbers of death group and the non-death group were very imbalanced (78 death cases and 725 non-death cases). 31 SNVs were identified to be significantly enriched in the death group (Figure 3D), although much fewer genomes carrying these SNVs. The top three SNVs, one non-synonymous mutation G16078A (nsp12: V880I) and two synonymous ones, G10396T on nsp5 and G21207T on nsp16, were detected in both the death and non-death group with an odds ratio of 3.66, 28.96, and 14.46, respectively. All other SNVs were observed only in very few numbers of the death genomes. The imbalance between death and non-death sample sizes was observed in Italy as well (15 vs 458, Figure 3E). But unlike India, 10 SNVs significantly enriched in the death group also presented much higher occurrence ratios in the group, although only 15 death-related genomes were collected in Italy. For example, the top two SNVs C7639T (nsp3: F1640F) and G20398A (nsp15: E260K) were identified in 40.0% and 20.0% of death cases compared to only 7.42% and 0.87% of non-death cases, respectively. Very few genomes harbored both SNVs simultaneously, suggesting the independence or mutual exclusion of SNVs C7639T and G20398A. Surprisingly, none of SNVs identified above was observed among any two of these five countries (Figure 3F).

Next, we employed the elastic-net regularized logistic regression model to evaluate the relationship between the mortality status and significant SNVs (*p* <0.01 and FC > 2) mentioned above in Brazil. The elastic-net regularized logistic regression model is well known to deal with sparse data as well as colinear of covariates by combining L1 and L2 regularization (14, 15). With 5-fold cross-validation, 4 out of 12 SNVs, G25088T, T25A, and two SNVs close to 3’end of the viral genome, G29861T and G29864A, had significant coefficients which can be used in the model to predict mortality status in Brazil as below,

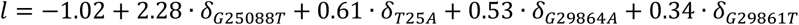

where *l* is log-odds and

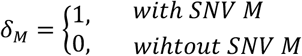

The model reached to the area under the curve (AUC) of 0.84. It is interesting that the second top SNV (Figure 3A), C12053T, was not included in the model, because C12053T occurred simultaneously with G25088T in the genomes in Brazil, while G25088T prevailed in the relationship with mortality status.

### Protein structure variations due to SNVs

Four SNVs in groups A.E1-A.E3 are non-synonymous variants, including ORF10:V30L (G29645T), N: A220V (C28923T), and two on S protein, A222V (C22227T) and L18F (C21614T) (Figure 1A). The structure modeling derived that all of them were on the surface area of corresponding proteins. Through mutagenesis analysis, the changes in the AAs at these sites were all incurring clashes illustrated by red disks with nearby residues (Figure 4A-D), with potential impacts on protein configuration. For example, N: A220V was located at the bottom of a pocket. Mutation to Valine resulted in a more hydrophobic state which may affect potential binding activity on the site. Such changes of protein conformation may also affect virus pathogenesis or vaccine response through interacting proteins.

**Figure 4.**
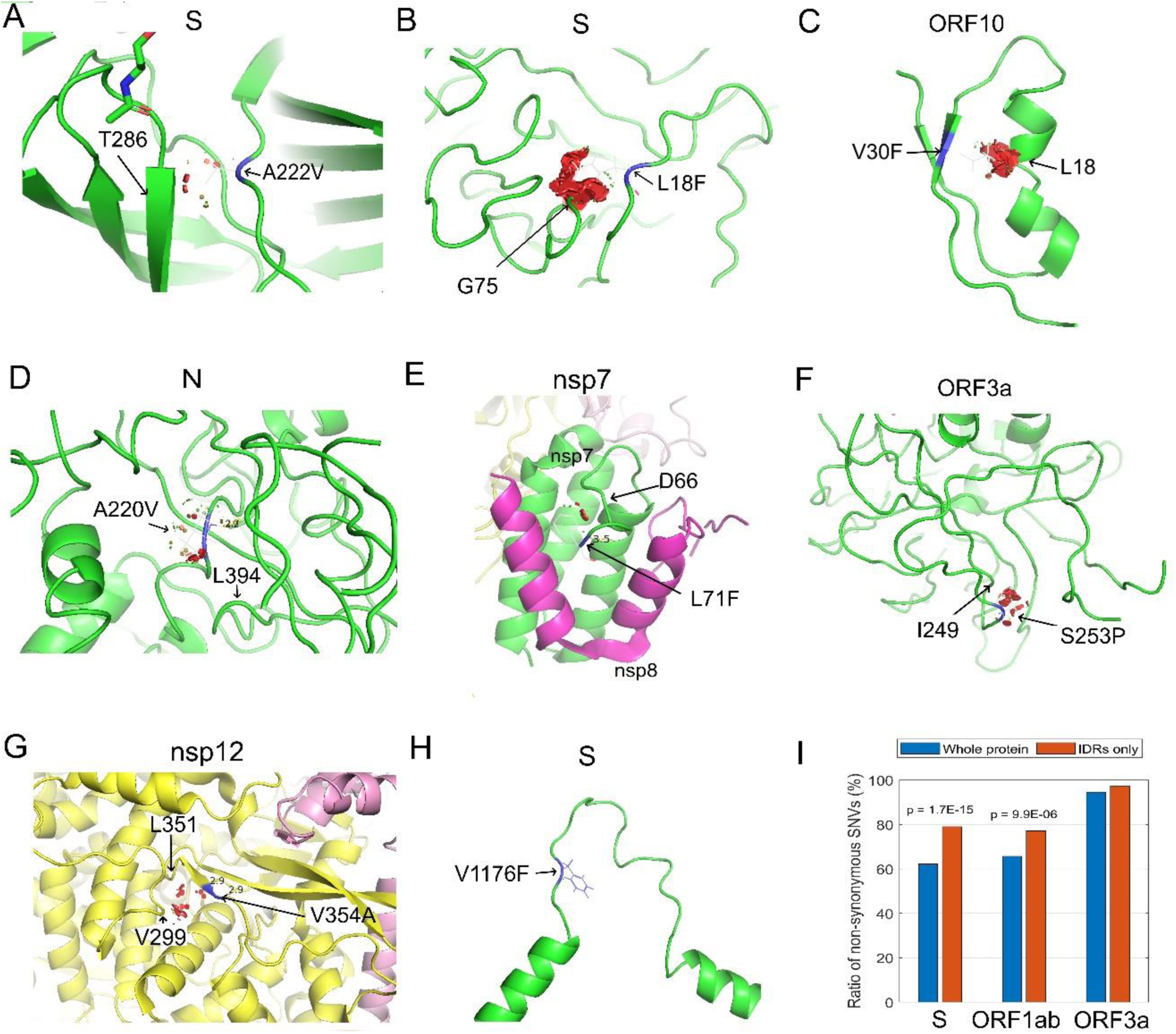
Protein structure variation caused by selected non-synonymous SNVs. (A) S: A222V. (B) S: L18F. (C) ORF10: V30F. (D) N: A220V. (E) nsp7: L71F. (F) ORF3a: S253P. (G) nsp12: v354A. (H) S V1176F. (I) Ratios of non-synonymous SNVs in the whole or IDRs region of proteins, S, ORF1ab, and ORF3a.

Similarly, some of the non-synonymous SNVs significantly over-represented in the death group in Brazil also caused clashes with nearby residues (Figure 4E-H). The mutation on nsp7: L71F (C12053T) might clash with D66, while ORF3a: S253P (T26149C) with I249, nsp12: V354A (T14501C) with V299 and L351, and N: T205I (C28887T) with A414. These variants also occurred on protein surface area. Consequently, the clashes may affect their pathogenesis or vaccine response, suggesting contributions to higher mortality rate. Interestingly, mutation S: V1176F (G25088T) was in the loop structure of two α-helix in the C-terminal of S, causing increased mass. This may have impacts on the protein structure which is necessary for virus entry into the host cell.

Intrinsic disordered region (IDR) in a protein represents an unfixed or disordered three-dimensional structure due to lack of sufficient hydrophobic AAs (31, 32). The AA composition may influence the IDR conformational state, further resulting in variations of functional elements within IDRs and protein function alteration. IDRs determined from experiments cover 28.0% of S protein sequence, 27.6% of ORF3a protein sequence, and 3.8% of ORF1ab polyprotein respectively (31, 32). Non-synonymous SNVs were found on approximately 62.3%, 65.7%, and 94.6% of S, ORF1ab and ORF3a, respectively (Figure 4I).

Significantly higher SNV occurrence ratios were observed in IDRs of S (79.0%, *p* = 1.7×10^−15^) and ORF1ab (77.1%, *p* = 9.9×10^−6^), but not in IDR of ORF3a (97.4%, *p* = 0.12). For instance, G22992A (S: S477N) was in S protein receptor-binding domain (RBD), which is a flexible and disordered loop in the unbound state but later becomes ordered in all the available ACE2-bound SARS-CoV-2 S structures (33). The decreased hydrophobicity and increased mass upon mutation from S to N may affect the flexibility of this region and the binding of S protein to ACE2. Another mutation C21614T (S: L18F) in emerging group A.E3 SNV is located in N-linked glycan sites, which likely play a role in protein folding and immune evasion and may have implications in viral virulence and vaccine design (34). Other mutations in IDRs of S, e.g., C21575T (L5F), G25049T (D1163Y), G25062T (G1167V), are located nearby N-linked glycan sites (11, 1, and 5 residues, respectively). They may have impacts on glycosylation which in turn play a role in protein folding and immune evasion.

## Discussion

In this study, we monitored group structure changes of SARS-CoV-2 SNVs over time by comparing clustering results before and after June 2020. The representative SNVs in group A (5) had become dominant by covering over 99% of detected SARS-CoV-2 genomes since June 2020, while two other groups (B and D) were detected in less than 0.3% of the population. The occurrence ratio of group C SNVs also reduced compared to the previous result before June 2020 (5). While the majority of groups A and C SNVs were observed as mutually exclusive with each other before June 2020, 95% of genomes harboring the representative SNV in the group C, G11083T (ORF1ab: L3606F), also carried group A SNVs after June 2020, confirming the ruling role of group A in current SARS-CoV-2 genomes. Several emerging SNVs, e.g., groups A.E1-E3, co-occurred with group A representative SNV, S: D614G, in United Kingdom and other Northern European countries (Figures 1B-1C). The occurrence ratios of these subgroup SNVs had increased quickly since August 2020. For example, group A.E1 SNVs, represented by C22227T (S: A222V), G29645T (ORF10:V30L), and C28932T (N: A220V), increased from 10% in August to 70% in November of all samples in the same month, while the occurrence ratios of group A.E2, C27944T and G204T, and group A.E3, C21614T (S: L18F) and C24334T, increased to about 60% and 50%, respectively, during the same time period The results indicate that these SNV groups might make virus more contiguous. Our systematic study revealed that apart from the A.E1-3, more SNV groups occurred more than 3% of the SARS-CoV-2 genomes with distinct temporal patterns, even though some of them did not clearly show increasing trend over time (Figure 1). Cautions should be taken to monitor these SNVs with collectively dynamic changes.

In December 2020, a set of 23 changes or mutations (VUI-202012/01) were found to possibly drive infections in United Kingdom (35, 36). The set of signature variants includes 8 changes from S protein: deletion 69-70, deletion 144-145, N501Y (A23063T), A570D (C23271A), D614G, P681H (C23604A), T716I (C23709T), S982A (T24506G), D1118H (G24914C). Virus with these mutations were reported to be up to 70% more transmissible than previous strains, although there was “considerable uncertainty” and “no evidence” that these virus were more lethal or could render vaccines and treatments useless (37). These SNVs occurred independently before November 2020 without causing significant higher viral transmission. This might suggest that the collective mutations from these SNVs may speed up COVID-19 transmission. However, from the samples downloaded from GISAID, we did not observe high occurrence rates of these SNVs. We will keep track of them and try to understand the links between them and group E SNVs.

With integration of the clinical information, 30 SNVs were identified as significantly over-represented in the group associated with COVID-19 related death. The enrichment analysis as well as logistic regression model uncovered several SNVs as mortality-correlated for individual countries. For instance, G25088T and several other SNVs were observed significantly enriched in death cases in Brail. It can be adopted in a logistic regression model together with T25A, G29861T, and G29864A to predict the mortality status in Brazil. Regardless of potential biased sampling from limited genomes with clear clinical information, none of these mortality related SNVs were associated with any of two countries concomitantly. In other words, the SNVs related to mortality status were specific to the country, suggesting that it was not only SNVs themselves but also other unknown geographical features, e.g., potential environmental and human health factors, can contribute to COVID-19 lethality.

The protein structure analysis of emerging SNVs in groups A.E1-3 and selected mortality-associated SNVs showed that all of these mutations occurred on protein surface area. Some clashes introduced upon mutation may contribute to the higher level of transmission and even mortality rate. Further investigation of IDRs on S and ORF1ab protein showed that non-synonymous SNVs tended to appear in IDRs, suggesting the connections between IRDs of S and ORF1ab and their protein conformation and functions. More in-depth study to understand these effects may help therapeutic intervention and vaccine design.

## Data Availability

Yes. The data/results will be available upon requests.

## Acknowledgement

This work was partially supported by the National Institutes of Health (P30CA082709 to J.W.), the Walther Cancer Foundation, and Center for Computational Biology and Bioinformatics (CCBB) at Indiana University School of Medicine (Pilot grant to L.Y. and J.W.). Special thanks to researchers for depositing whole genomic sequences of Novel Pneumonia Coronavirus (SARS-CoV-2/hCoV-19/2019-nCoV) at the Global Initiative on Sharing All Influenza Data (GISAID) EpiFluTM. We are thankful for the technical support from the GISAID in downloading the SARS-CoV-2 genomes. We appreciate helpful discussion with Dr. Yong Zang at Indiana University School of Medicine.

